# Characterizing Longitudinal Antibody Responses in Recovered Individuals Following COVID-19 Infection and Single-Dose Vaccination in British Columbia, Canada: a Prospective Cohort Study

**DOI:** 10.1101/2022.09.28.22280429

**Authors:** Andrea D. Olmstead, Aidan M. Nikiforuk, Sydney Schwartz, Ana Citlali Márquez, Tahereh Valadbeigy, Eri Flores, Monika Saran, David M. Goldfarb, Althea Hayden, Shazia Masud, Agatha N. Jassem, Muhammad Morshed, Inna Sekirov

## Abstract

**Background:** Investigating antibody titres in individuals who have been both naturally infected with SARS-CoV-2 and vaccinated can provide insight into antibody dynamics and correlates of protection over time.

**Methods:** Human coronavirus (HCoV) IgG antibodies were measured longitudinally in a prospective cohort of PCR-confirmed, COVID-19 recovered individuals (k=57) in British Columbia pre- and post-vaccination. SARS-CoV-2 and endemic HCoV antibodies were measured in serum collected between Nov. 2020 and Sept. 2021 (n=341). Primary analysis used a linear mixed-effects model to understand the effect of single dose vaccination on antibody concentrations adjusting for biological sex, age, time from infection and vaccination. Secondary analysis investigated the cumulative incidence of high SARS-CoV-2 anti-spike IgG seroreactivity equal to or greater than 5.5 log10 AU/mL up to 105 days post-vaccination. No re-infections were detected in vaccinated participants, post-vaccination by qRT-PCR performed on self-collected nasopharyngeal specimens.

**Results:** Bivariate analysis (complete data for 42 participants, 270 samples over 472 days) found SARS-CoV-2 spike and RBD antibodies increased 14-56 days post-vaccination (p<0.001) and vaccination prevented waning (B=1.66 [95%CI: 1.45-3.46]); while decline of nucleocapsid antibodies over time was observed (B=-0.24 [95%CI: -1.2-(−0.12)]). A non-significant trend towards higher spike antibodies against endemic beta-HCoVs was also noted. On average, SARS-CoV-2 anti-spike IgG concentration increased in participants who received one vaccine dose by 2.06 log10 AU/mL (95%CI: 1.45-3.46) adjusting for age, biological sex, and time. Cumulative incidence of high SARS-CoV-2 spike antibodies (>5.5 log10 AU/mL) was 83% greater in vaccinated compared to unvaccinated individuals.

**Conclusions:** Our study confirms that vaccination post-SARS-CoV-2 infection provides multiple benefits, such as increasing anti-spike IgG titers and preventing decay up to 85 days post-vaccination.

## 1. Introduction

The coronavirus disease 2019 (COVID-19) pandemic, caused by the novel beta (β)-coronavirus, severe acute respiratory syndrome coronavirus 2 (SARS-CoV-2), has caused significant morbidity, mortality, economic impact, and disruption of health care and societal systems. Prior to the emergence of COVID-19, four seasonal human coronaviruses (HCoV) were identified that typically cause self-limited respiratory infections with mild symptoms, i.e., the ‘common cold’ (1). Like SARS-CoV-2, HCoV-OC43, and HCoV-HKU1 are β-coronaviruses, while HCoV-229E and HCoV-NL63 and classified as α-coronaviruses (2). Coronavirus genera are separated by unique serological and genomic characteristics; viral species from the same genus share cross-neutralizing (non-specific) antibodies which arise from homology in viral genes and structural proteins (3).

In the province of British Columbia (BC), Canada, the first confirmed case of COVID-19 was reported on January 25, 2020; strict and swift public health measures were largely effective at controlling spread during the first wave, which peaked locally between the third week of March and late April in 2020 (4). During the first epidemiological wave of the pandemic, little was known about antibody responses to SARS-CoV-2 infection and studies were needed to understand if and how quickly infected individuals develop a detectable, protective, and durable antibody-mediated immune response. Understanding the durability or waning of antibodies over time helps elucidate the risk of re-infection and inform vaccination schedules. Studies have shown that most SARS-CoV-2 infected individuals seroconvert within 14-28 days; the spike (S) and the nucleocapsid (N) proteins elicit the strongest humoral response (5, 6). Predictably, SARS-CoV-2 antibody concentrations wane over time; the rate of decline varies widely depending on various factors (e.g., age, biological sex, and disease severity) (7–9). Neutralizing antibodies acquired naturally or from vaccination protect against infection and re-infection (10). Several studies have shown a strong correlation between anti-S, anti-RBD and neutralizing antibody titers, as such measuring anti-S and anti-RBD can be used as a proxy for antibody-mediated protection (11–13).

Despite the success of SARS-CoV-2 vaccines, many individuals are still hesitant to be immunized against COVID-19; supply shortages combined with social and economic inequity hamper global vaccination efforts (14–16). The study of antibody dynamics following natural infection and the impact of vaccination on those who have been previously infected is needed, as novel SARS-CoV-2 variants with increasing capacity to escape pre-existing immunity continue to evolve and spread (17–19). Observational studies agree that vaccination benefits those who have been previously infected, but the number of doses required for optimal protection remains unclear (20–22).

We describe a prospective cohort that was established to monitor antibody responses over three months in people that recovered from SARS-CoV-2 infection. Many participants were offered a single dose COVID-19 vaccine during the study; therefore, we expanded the aims to study the dynamics of antibodies against both SARS-CoV-2, as well as endemic HCoVs, in recovered individuals pre- and post-vaccination against SARS-CoV-2, and investigated their relationship with age, biological sex, and symptom duration.

## 2. Materials and Methods

### 2.1. Study Design

A prospective observational cohort termed CARE (Characterizing the Antibody Response to Emerging COVID-19) was established from individuals who recovered from SARS-CoV-2 infection, for the purposes of investigating antibody responses against several SARS-CoV-2 epitopes (full spike (S), receptor binding domain (RBD) and nucleocapsid (N)), as well as against the S protein of endemic HCoVs (OC43, HKU1, NL63, 229E), at least 2 weeks post natural SARS-CoV-2 infection, with or without subsequent SARS-CoV-2 vaccination. Information on age, biological sex and date of PCR-diagnosis was collected through medical records, while information on the duration of COVID-19 symptoms, hospitalization, and vaccination was collected through an online self-reporting survey. The date of confirmed PCR diagnosis of SARS-CoV-2 infection was used to estimate *‘*days post-infection’. Survey data and participant informed consent were collected and managed using REDCap electronic data capture tools hosted at BC Children’s Hospital (Vancouver, BC). REDCap (Research Electronic Data Capture) is a secure, web-based application designed to support data capture for research studies (23). Participants were enrolled in the cohort from November 19^th^, 2020, to September 7^th^, 2021. During this time the most prevalent SARS-CoV-2 variant in British Columbia transitioned between the Alpha, Beta, Gamma and Delta genotypes. The Beta variant was detected in late 2020 and January 2021, increase prevalence of the Alpha variant shortly followed and it remained dominant until June 2020. The Gamma variant was first detected in late February 2021, it’s incidence surpassed Alpha in July 2021. Public health surveillance first recorded the Delta variant in March, and it was responsible for most sequenced cases over the summer from July to September 2021(24). All data analysis was performed in R version 4.0.4 using the packages: ‘DataExplorer’, ‘survival’, ‘survminer’, ‘dplyr’, ‘ggfortify’, ‘tableone’, ‘naniar’, ‘RColorBrewer’, ‘lme4’, ‘mgcv’, ‘gam.check’ and ‘readr’ (25).

### 2.2. Recruitment Criteria

Adults 18 years of age and older from the greater Vancouver metropolitan area were recruited if they had a confirmed PCR-positive SARS-CoV-2 infection and if they were no longer required to self-isolate per the BC provincial public health guidelines (i.e., tested positive for SARS-CoV-2 at least 14 days prior). The study protocol was approved by the University of British Columbia (UBC) Clinical Research Ethics Board (H20-01089).

### 2.3. Sample Collection and Processing

Participants were asked to donate blood samples collected by venipuncture (for serological testing) and concurrent self-collected saline gargle samples (for SARS-CoV-2 real-time-polymerase chain reaction (PCR) testing), every two weeks for 3 months post-recruitment (up to 7 collections total). Blood was drawn in gold-top serum separator tube with polymer gel (BD, cat# 367989); after at least 30 minutes of clotting at room temperature, the blood sample was then centrifuged at 1,400 G by staff at the collection site and sent to the British Columbia Centre for Disease Control Public Health Laboratory (BCCDC PHL). At the BCCDC PHL the samples were divided into serum aliquots that were frozen at −80°C within four hours of receipt. Blood collections occurred at four sites in the Greater Vancouver Area, British Columbia Canada: BC Children’s Hospital, St. Paul’s Hospital, Abbotsford Regional Hospital and Surrey Memorial Hospital. Saline gargle samples were self-collected by the participants at home, in accordance with well-validated instructions (26), on the day of blood collection and transported to the BCCDC PHL by the blood collection site. Self-collected saline gargle samples were tested for SARS-CoV-2 by PCR. SARS-CoV-2 whole genome sequencing was done on a subset of participants’ SARS-CoV-2 positive diagnostic clinical specimens (27). All molecular, genomic, and serological testing (described below) for participant specimens was conducted centrally at the BCCDC PHL.

### 2.4. Measurements of Humoral Immunity

All serum samples were initially tested using a combination of three Health Canada approved chemiluminescent immunoassays: 1) total antibodies to SARS-CoV-2 RBD (Siemens SARS-CoV-2 Total Assay [COV2T]), 2) total antibodies to SARS-CoV-2 S (Ortho VITROS™ Anti-SARS-CoV-2 Total) and IgG anti-N antibodies (Abbott ARCHITECT™ SARS-CoV-2 IgG), as per manufacturer guidelines, with results interpreted as reactive or non-reactive using the manufacturer-recommended signal to cut-off ratios (28, 29). All available samples were then tested using the V-PLEX COVID-19 Coronavirus Panel 2 (IgG) (Mesoscale Diagnostics LLC (MSD): #K15369U), the diagnostic accuracy of the MSD assay was previously validated through comparison with alternative Health Canada approved tests at the BCCDC PHL (30). The MSD assay provides quantitative measures of IgG antibodies against RBD, S and N SARS-CoV-2 epitopes, as well as IgG antibodies against S of the four seasonal HCoVs. Serological specimens were processed as previously reported (30). Quantitative antibody levels expressed as log10 antibody units (AU)/ml were recorded and evaluated for all tested samples. MSD results were interpreted as reactive or non-reactive using the MSD recommended signal thresholds for serum: SARS-CoV-2 anti-S IgG = 1960; anti-N IgG = 5000; anti-RBD IgG = 538. Cutoffs (derived at the BCCDC PHL) for seasonal HCoVs seropositive status are as follows: HCoV-229E anti-S IgG = 1700; HCoV-HKU1 anti-S IgG = 900; HCoV-NL63 anti-S IgG = 270; HCoV-OC43 anti-S IgG = 2000 (31). Samples were stratified by collection time to < 6 months and ≥ 6 months post infection and percent positivity was compared using a Chi-square test (χ²test).

### 2.5. Power Analysis for Investigating Association between IgG Concentration and Vaccination

A power calculation was conducted to determine the minimum number of paired participant samples needed to estimate at least a 70% association between COVID-19 vaccination and HCoV anti-IgG antibody concentration. Antibody concentrations were assumed to be normally distributed with a standard deviation of one. A significance level of 5% and two-sided alternative were used (32).

### 2.6. Analytic Data Selection

To analyze antibody dynamics, an analytic dataset was selected from the CARE COVID-19 cohort. At least k = 18 paired participant samples are required to estimate a 70% or greater association between COVID-19 vaccination and anti-HCoV IgG antibody concentration. Exclusion criteria were applied to select an analytic dataset from k = 57 participants with n = 341 observations. One participant had no follow up samples and was omitted from the analytic dataset (k = 1, n = 1). Six participants were excluded because they were vaccinated before collection of their baseline sample (k = 6, n = 37). Eight participants were removed from the analytic dataset because of missing data in their survey results (k = 8, n = 33). After applying the exclusion criteria, the analytic dataset contained k = 42 participants with n = 270 observations (Figure S1). There were k= 41 participants with >1 pre-vaccine sample (n=210 pre-vaccine observations) used for analysis of antibody waning pre-vaccination.

### 2.7. Bivariate Data Analysis

#### 2.7.1 Antibody Waning

Waning of SARS-CoV-2 antibodies prior to vaccination was investigated in independent participant specimens measured at baseline using linear regression. HCoV anti-IgG antibody signals at baseline were compared to signals 14-56 days post-vaccination using a paired t-test in a sample of n=21 participants who received a COVID-19 vaccine during the study. Waning of SARS-COV-2 specific IgG was measured prior to vaccination between participants using linear regression. Participant’s baseline samples (defined as the first specimen taken after enrolling in the study) were plotted for anti-S and anti-N IgG over time.

#### 2.7.2 Descriptive statistics

Bivariate analysis was conducted between the exposure (a single dose of a Health Canada approved COVID-19 vaccine) and outcome (SARS-CoV-2 anti-S or anti-N IgG) of interest at baseline. Baseline represents the time of a participant’s first blood draw after enrollment. The bivariate relationship between vaccine status and covariates was examined by t-test or chi square test depending on variable type. HCoV anti-IgG antibody signals were transformed to the logarithmic base ten scale for conformation to normality and ease of interpretation.

### 2.8. Primary Analysis

Primary analysis used a multivariable linear mixed-effects model to regress SARS-CoV-2 anti-IgG concentration on vaccine status adjusting for dependency within participant samples and covariates defined as potential confounders by the common cause criterion (33, 34). Separate models were fit for anti-S and anti-N IgG signals. Unconditional mean models were used to find the intraclass correlation coefficient (ICC) before covariates were added to build fixed effect models (35). Effect modification terms were assessed by the Akaike information criterion and included in the fixed effect models to understand if time from vaccination influences SARS-CoV-2 antibody concentrations (36).

### 2.9 Secondary Analysis

Secondary analysis employed a Kaplan-Meier curve to estimate the cumulative incidence of seroreactivity stratified by vaccine status. The survival function was transformed to cumulative incidence by 1-S(τ) (37). Seroreactivity was defined from the distribution of SARS-CoV-2 anti-S IgG concentration at first blood draw (baseline); the 95^th^ percentile was chosen as the threshold (5.5 log10 AU/ml). Participants were censored if they were not seroreactive before loss to follow-up (right censoring). A log-rank test was used to test the hypothesis that the cumulative incidence of seroreactivity between unvaccinated and vaccinated persons, who have been previously naturally infected with SARS-CoV-2, does not differ (38).

## 3. Results

### 3.1. CARE COVID-19 Cohort

Fifty-seven individuals with COVID-19 were recruited through the CARE COVID-19 Cohort. Subjects (17 male, 40 female; 18 to 76 years old) represented a range of COVID-19 disease severity cases. Most subjects had a mild case of COVID-19, defined as not requiring hospitalization; 6 reported being asymptomatic and 12 reported experiencing fever. Only four subjects (7%) reported being hospitalized for COVID-19; one required intensive care. The observed case severity distribution was consistent with the general distribution of COVID-19 disease severity in BC (∼5% of diagnosed cases hospitalized as of April 2022) (39).

Participants were required to have recovered from COVID-19 (i.e., 10 days post-PCR diagnosis) before providing their first blood and saline gargle sample. Collection dates ranged from 18-490 days (median 152 days) since a positive PCR test (used as proxy for time since infection), with the baseline collection date ranging from 18-339 days (median 114 days). Participants submitted between 1 and 7 samples, with approximately 2 weeks (median 14 days; range 7–83 days) between each collection, with an average of 6 samples collected per participant and a total of 341 samples collected. No reinfections or persistent virus shedding were detected in self-collected saline gargle samples using RT-PCR (data not shown).

Virus whole genome sequencing was performed (27) on diagnostic samples from 14 participant specimens that were available for analysis, to determine the SARS-CoV-2 variant responsible for infection. SARS-CoV-2 variants were classified as ancestral (e.g., A.1) (n=4) and the D614G mutant (e.g., B.1) (n=10), which is consistent with variants circulating at the time of respective participants’ diagnoses (40). Whole genome sequencing data was missing for ∼71% of participants and; therefore, not included as a covariate in the analysis. Multiple studies corroborate no significant difference in neutralising antibodies between the alpha variant and the ancestral isolate post mRNA vaccination with BNT162b2 or mRNA-1273. Noteworthy reduction of post-vaccination neutralising sera was observed for the beta variant in persons vaccinated with mRNA-1273(41).

### 3.2 Comparison of anti-SARS-CoV IgG Antibody Responses Across Four Commercial Assays

All available samples (n=340; 1 missing) were initially tested using a combination of three commercial serology assays supplied by Siemens (COV2T), Abbott (ARCHITECT™), or Ortho (VITROS™) clinical diagnostics. Of 340 samples tested, 338 were classified as reactive using at least one assay (Table S1). All available samples (n=339) were subsequently tested using a highly sensitive and multiplex electro chemiluminescent assay offered by Meso Scale Diagnostics (MSD). Percent positivity differed across the platforms and by antigenic target. Overall detection of anti-S was more sensitive than anti-N SARS-CoV-2 IgG. Comparing anti-S results, the Ortho assay had the highest positivity rate (100%) followed by Siemens (95%) and MSD (89%) (Table S1). For anti-N results MSD (58%) outperformed Abbott (47%) with a 11% increase in positivity (χ²test, P=0.01). When samples were stratified by collection time to less than or greater than 6 months post-infection, the anti-N positivity rate decreased for both the Abbott (72% to 13%) and MSD, (76% to 33%) (P<0.001). A 7% decline in positivity was observed for anti-S (P=0.06) and 2% for anti-RBD (P=0.53) when tested by MSD (Table S1). Only antibody measurements from the MSD assay were used in the multivariable analysis as the anti-S IgG results compared well with Ortho and anti-N IgG results were superior to Abbott.

Waning of anti-S and anti-N IgG concentrations over time were measured between participants using the first baseline observation for each of the k=42 participants in the analytic dataset. Using linear regression analysis, overall waning was observed in both anti-N and anti-S and the slope did not differ significantly across the two measures (P = 0.46; Figure 1). On average SARS-CoV-2 antibodies wane at a rate of -0.0029 log10 AU/mL per day (P <0.001) or ∼ 4228 AU/mL per month. These results confirm waning of anti-SARS-CoV-2 antibodies over time in people who have recovered from natural SARS-CoV-2 infection before vaccination. Estimates of anti-ARS-CoV-2 IgG waning are calculated post-vaccination using a mixed-effects linear regression model and reported as the ‘primary analysis’.

**Figure 1.**
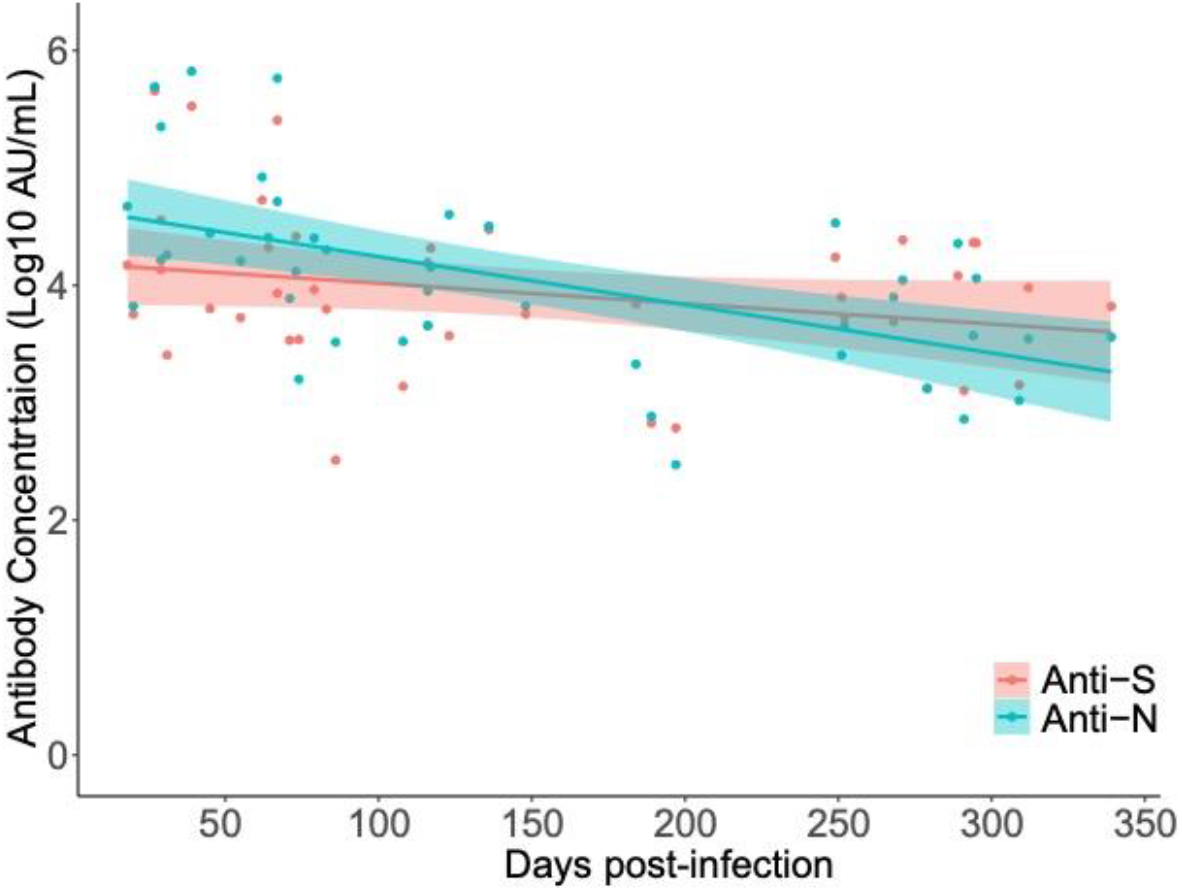
Longitudinal decay of SARS-CoV-2 anti-N and anti-S IgG concentration over time in natural SARS-CoV-2 infected CARE participants prior to vaccination (k= 42, n= 42 samples). Participant samples were restricted to the first collection date (baseline) and plotted independently. Linear regression was used to estimate the decrease in anti-S and anti-N titre over time since PCR test result.

### 3.3 Serological Response to SARS-CoV-2 Vaccination

Bivariate analysis was conducted on the analytic dataset to compare participant antibody responses pre- and post-vaccination for COVID-19. Participant’s serology results and survey responses are summarized and stratified at baseline by the exposure of interest, one dose of a COVID-19 vaccine (Table 1). No difference in the distribution of covariates between participants who received and did not receive a COVID-19 vaccine over the study period was observed for all variables except the number of participant visits. Though follow-up time did not significantly differ between the two groups, on average unvaccinated participants were observed 0.95 (approximately one) fewer times than those who received a COVID-19 vaccine (P=0.014) (Table 1). Importantly, age, biological sex, days from positive PCR test (diagnosis), symptom duration and endemic anti-coronavirus IgG signals did not differ by exposure at baseline; therefore, we expect limited confounding from these covariates when estimating the association between COVID-19 vaccination and anti-SARS-CoV-2 IgG signals. Covariates, which met the definition of a confounder by the common cause criteria, were adjusted for in the primary analysis using a linear mixed effects model.

**Table 1.**
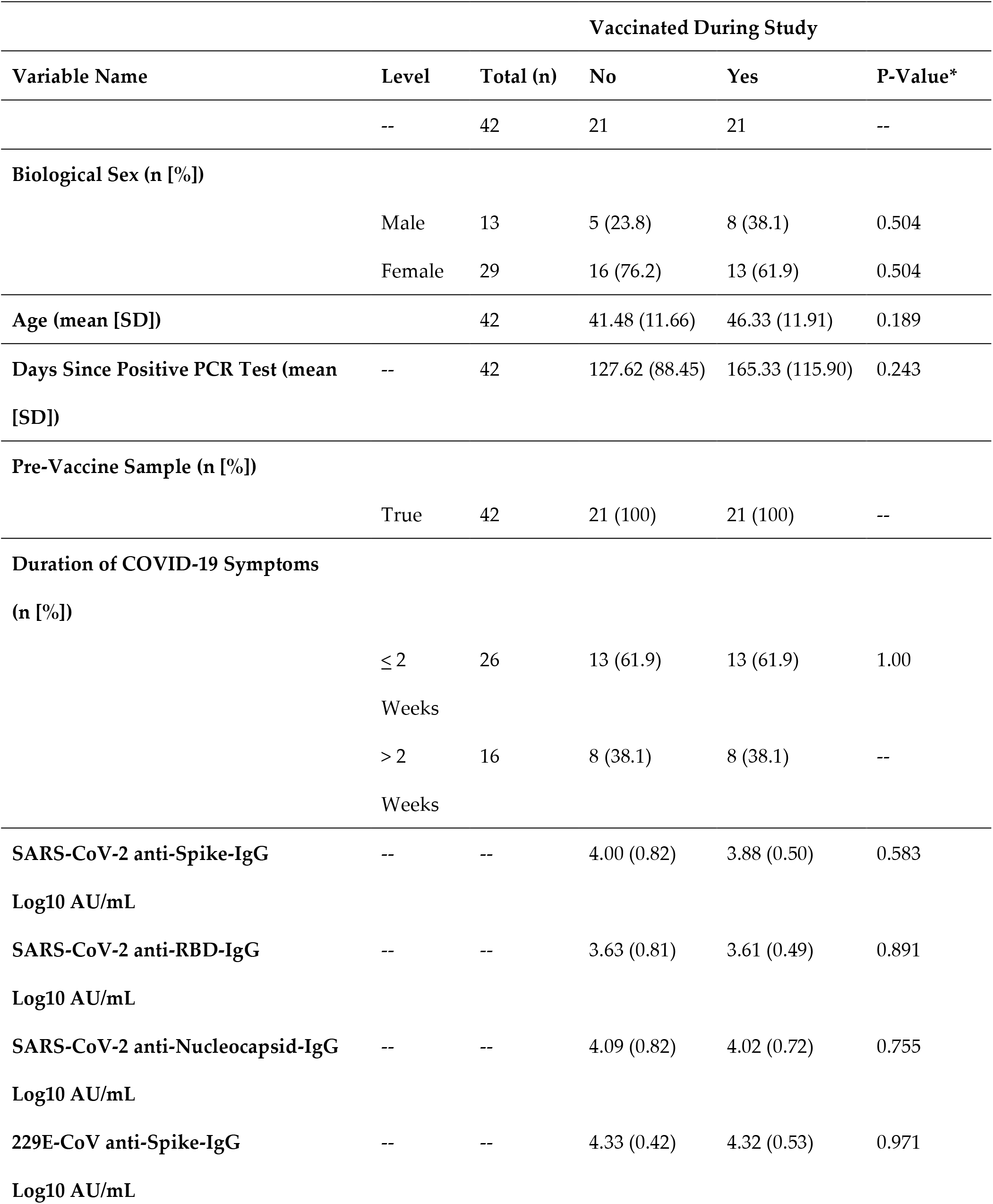
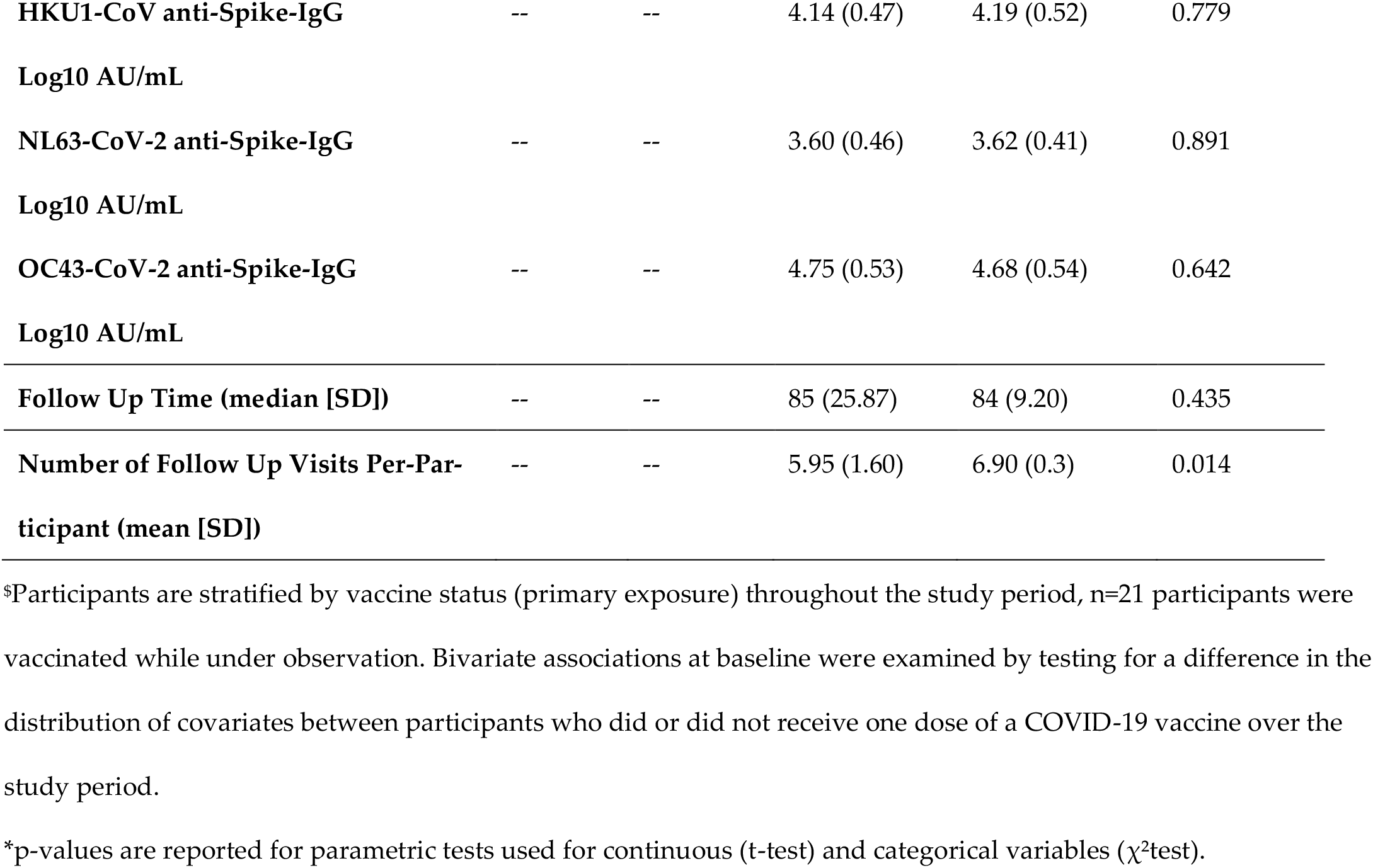
Descriptive statistics of study participants at the beginning of the study (baseline) in the analytic dataset with complete data (n=42) ^$^.

In k=21 paired participants, SARS-CoV-2 anti-S and anti-RBD IgG antibody concentrations increased post vaccination by 1.63 (P≤0.001) and 1.82 (P≤0.001) log10 AU/ml (Figure 2A & 2B). Anti-N antibody concentration continued to decrease post vaccination by -0.3 (P=0.03) log10 AU/ml (Figure 2C), consistent with waning observed prior to vaccination. Most participants (>99%) were found to be seropositive for anti-S antibodies against the endemic HCoVs. Post vaccination, anti-S antibody concentrations for endemic human -coronaviruses HCoV-HKU1 and HCoV-OC43 slightly increased (P=0.11 and P=0.07) (Figure 3B & 3D). No increase in antibody concentration was observed for the endemic human -coronaviruses HCoV-229E and HCoV-NL63 (P=0.43 and P=0.67) (Figure 3A & 3C).

**Figure 2.**
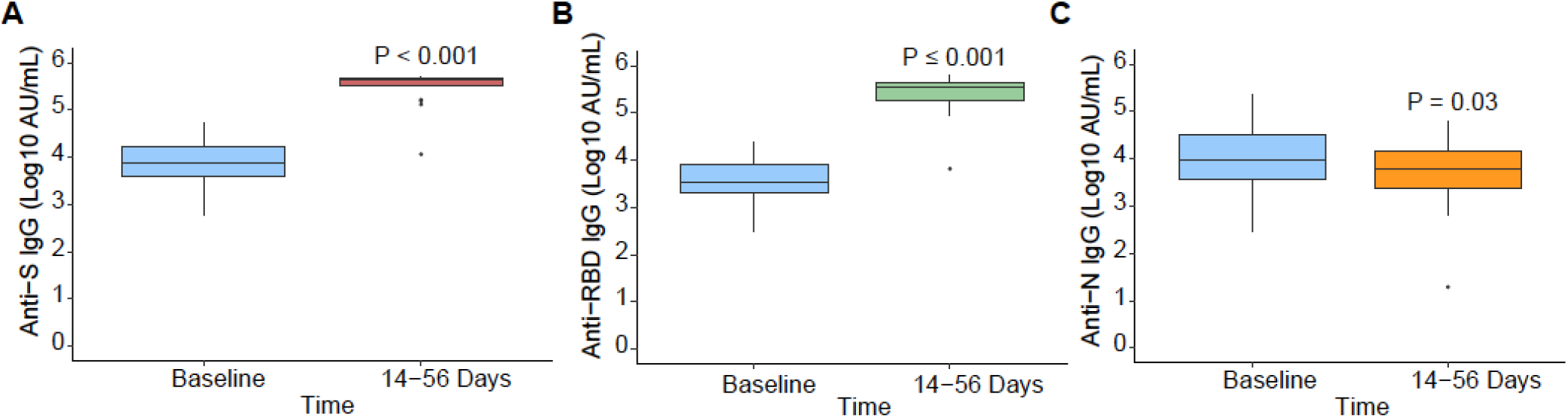
SARS-CoV-2 anti-IgG pre- and post-vaccination. Antibody signals in k=21 paired participants, who received a COVID-19 vaccine during the study, at baseline and 14 to 56 days post-vaccination, presented by individual SARS-CoV-2 antigen (k=21): **A)** anti-S, **B)** anti-RBD, **C)** anti-N. Differences in antibody signals were examined with a paired t-test.

**Figure 3.**
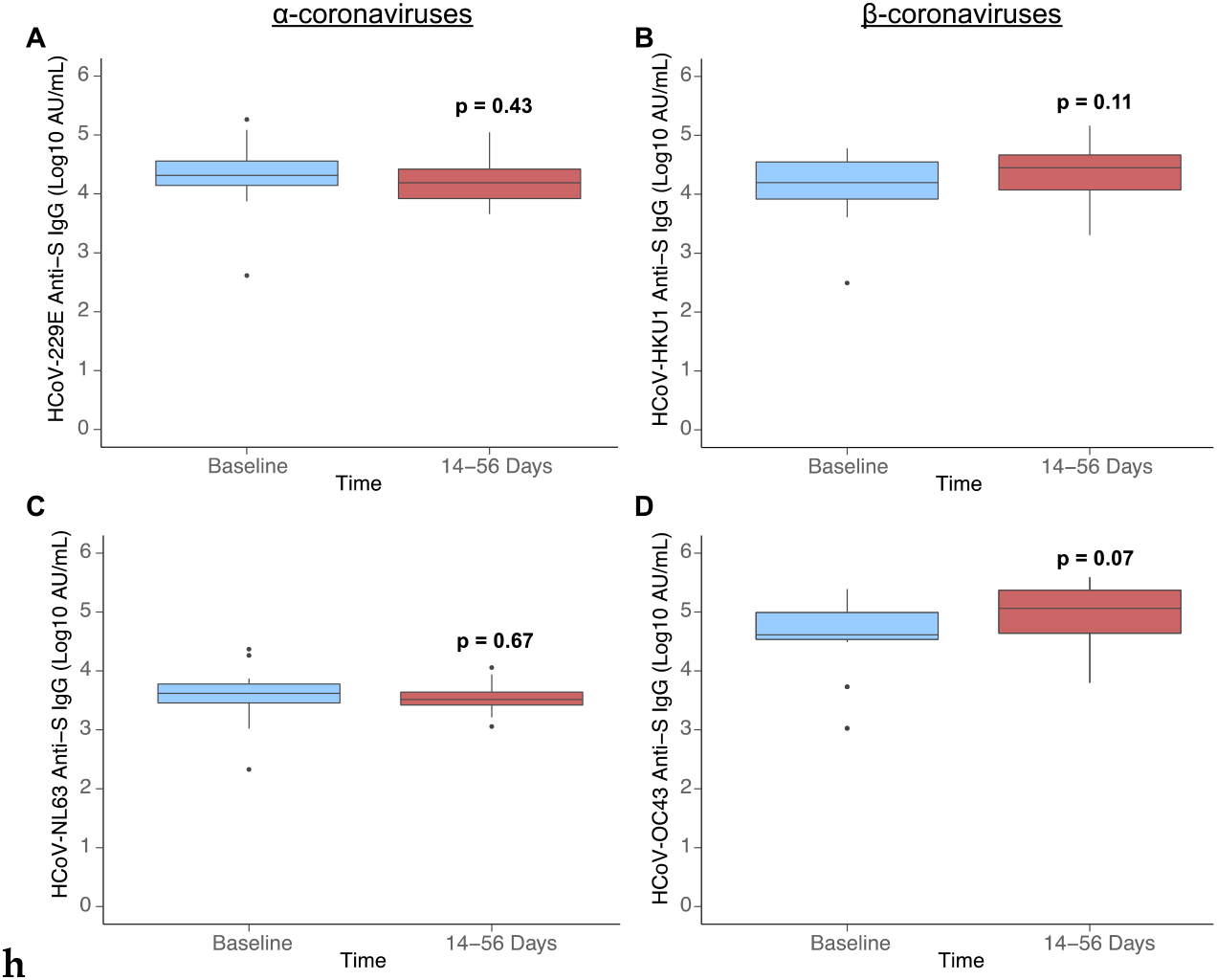
Endemic human coronavirus anti-S IgG antibody signals pre- and post-vaccination. HCoV antibody signals in n=21 paired participants, who received a COVID-19 vaccine during the study, measured at baseline (before vaccination) and 14 to 56 days post-vaccination, presented by HCoV species: **A)** HCoV-229E anti-Spike (S), **B)** HCoV-HKU1 anti-S, **C)** HCoV-NL63 anti-S, **D)** HCoV-OC43 anti-S. Difference in antibody signal was examined with a paired t-test.

### 3.4 Primary Analysis

Linear mixed-effects regression models were used to estimate intraclass correlation within participant samples and the relationship between COVID-19 vaccination and SARS-CoV-2 anti-S or anti-N IgG antibody concentration. An unconditional mean model was fit to partition within participant variation from between participant variation (Table 2). The minority of variation in SARS-CoV-2 anti-S IgG concentration was attributable to differences between participants (ICC=0.43) (Table 2). On average, anti-S IgG concentration increased over time in participants who received one dose of a COVID-19 vaccine during the study by 2.06 log10 AU/mL (95%CI: 1.45-3.46) adjusting for age, biological sex, days from positive PCR test (time) and effect modification between COVID-19 vaccination and time (Table 2). In the adjusted model, the ICC increased to 0.89 indicating that between participant differences (e.g., COVID-19 vaccination) explains most of the variation in SASRS-CoV-2 anti-S IgG antibody concentration. COVID-19 vaccination has a positive association with SARS-CoV-2 anti-S IgG antibody concentration, which increases over time. Variation in anti-N IgG concentration was due to differences between participants in the unconditional mean model (ICC=0.88). The average, anti-N IgG concentration decreased in vaccinated participants over time (−0.243 log10 AU/mL, 95%CI: -1.2 – [=0.12]) adjusting for age, biological sex, days from positive PCR test (time) and effect modification between COVID-19 vaccination and time (Table 2). Variation in anti-N IgG concentration after fitting the adjusted model was explained by within participant variance (ICC=30). Overall, these results indicate that waning of SARS-CoV-2 anti-N IgG is unaffected by COVID-19 vaccination. Anti-S IgG titers increase post vaccination; therefore, vaccination of recovered individuals benefits the durability of their humoral immune response.

**Table 2.** Summary of linear mixed effects models fit to examine the relationship between anti-S IgG log10 AU/ml (light grey) or anti-N IgG log10 AU/ml (dark grey) and COVID-19 vaccination status adjusting for: biological sex, age, and time from clinical diagnosis.

| Unconditional Mean Model (S) | Intraclass Correlation Coefficient |  |  |
| --- | --- | --- | --- |
| Participant ID (n=42) | 0.434 |  |  |
| Residual | 0.566 |  |  |
| Random Intercept Model | Variable | Fixed Effect Estimate | 95%CI |
| *Anti-Spike IgG* | Intercept | 4.84 | 3.27-6.39 |
|  | Vaccine-Yes | 0.40 | -0.41-1.20 |
|  | Biological Sex-Male | 0.93 | 0.068-1.79 |
|  | Age (Years) | -0.029 | -0.063-0.0057 |
|  | Time from +ve PCR Test<sup>a</sup> | -0.20 | -0.47-0.054 |
|  | Vaccine: Time<sup>a</sup> | 1.86 | 1.39-2.21 |
| Random Effects | Intraclass Correlation Coefficient |  |  |
|  | 0.893 |  |  |
| Unconditional Mean Model (N) |  |  |  |
| Participant ID (n=42) | 0.875 |  |  |
| Residual | 0.125 |  |  |
| Random Intercept Model | Variable | Fixed Effect Estimate | 95%CI |
| *Anti-Nucleocapsid IgG* | Intercept | 3.14 | 2.48-3.79 |
|  | Vaccine-Yes | -0.080 | -0.42-0.26 |
|  | Biological Sex-Male | 0.27 | -0.095-0.63 |
|  | Age (Years) | 0.016 | 0.0017-0.03 |
|  | Time from +ve PCR Test<sup>a</sup> | -0.40 | -0.53-(-0.27) |
|  | **Vaccine: Time<sup>a</sup>** | -0.077 | -0.25 – 0.11 |
| **Random Effects** | **Intraclass Correlation Coefficient** |  |  |
|  | 0.30 |  |  |

\*An effect modification term was incorporated to explore how the effect of vaccination on antibody concentration differs by time since diagnosis with a PCR test. Unconditional means models were fit to partition the variance by participant without inclusion of other exposure variables. Fixed effect models were built by applying the common cause criterion to select covariates which are a cause of the exposure, outcome, or both.

### 3.5 Secondary Analysis

Secondary analysis used the Kaplan-Meier method to estimate the cumulative incidence of seroreactivity above a defined threshold in vaccinated and unvaccinated participants over time. Seroreactive status was classified by the threshold of ≥ 5.5 log10 AU/mL SARS-CoV-2 anti-S IgG, as described in Methods. Participants with antibody measurements equal to or greater than the threshold were considered reactive. Over the 105 days follow up from baseline (first antibody measurement available for participants post-infection), 88% (95%CI: 42-98%) of vaccinated participants (n=16) were seroreactive compared to 5% (95%CI: 0-14%) of unvaccinated participants (n=1) (P= 0.03) (Figure 4). A single dose of COVID-19 vaccine increases the probability of a SARS-CoV-2 anti-S IgG antibody concentration ≥ 5.5 log10 AU/mL by 83%.

**Figure 4.**
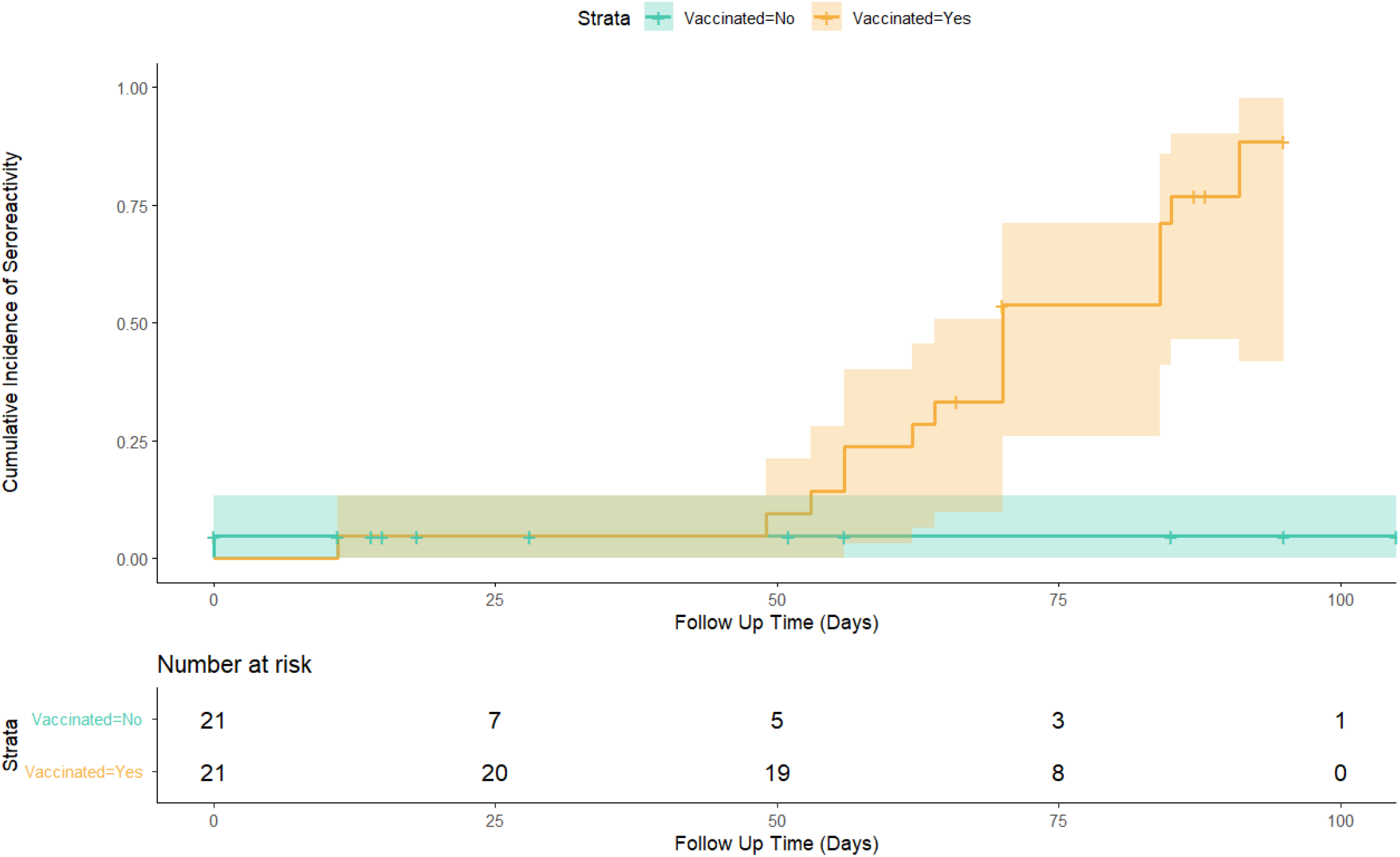
Cumulative incidence of seroreactivity (≥ 5.5 SARS-CoV-2 anti-S IgG Log10 AU/mL) days from participant’s first blood draw at baseline, stratified by vaccination status over the study period. Vaccinated participants achieved antibody titres not possible from natural infection alone (unvaccinated participants). Within 105 days of follow up, 88% (95%CI: 42-98%) of vaccinated participants were seropositive, an increase of 83% in comparison to the unvaccinated group (P= 0.03). In previously naturally infected individuals, COVID-19 vaccination increases SARS-CoV-2 anti-S IgG concentration over time to levels which are not attained by natural infection alone. No re-infections were detected by qRT-PCR in the vaccinated or unvaccinated group during the study period, specimens were self-collected.

## 4. Discussion

### 4.1. Summary of Results

A prospective cohort study was carried out in British Columbia to observe anti-SARS-CoV-2 and anti-endemic HCoV antibody dynamics in participants who were infected with SARS-CoV-2, a subset received the first dose of a Health Canada approved SARS-CoV-2 vaccine during the follow up. Several commercial serology assays were used to detect anti-Coronavirus antibodies; detection of anti-SARS-CoV-2 antibodies was confirmed in all available samples, although both anti-S and anti-N antibodies declines over time post-infection. Bivariate analysis found that vaccination significantly increased the titer of SARS-CoV-2 anti-S IgG antibodies 14-56 days post vaccination; no significant association was found between SARS-CoV-2 vaccination and endemic HCoV anti-S IgG antibodies, although antibodies against the S protein of beta-coronaviruses trended upwards, unlike those against the S protein of alpha-coronaviruses. Vaccination was not observed to boost SARS-CoV-2 anti-N IgG titers, which waned overtime in both vaccinated and unvaccinated individuals. The rate of anti-N waning was approximately double that of anti-S. Secondary analysis used a Kaplan-Meier model to estimate the cumulative incidence of anti-S antibody titers equal to or above 5.5 log10 AU/mL (‘seroreactivity threshold’) in those vaccinated and unvaccinated. In the vaccinated group, 88% (95%CI: 42-98%) of participants had SARS-CoV-2 anti-S IgG titers greater than or equal to this threshold, while this level was achieved in only one unvaccinated participant measured twenty-seven days post infection.

Despite overall antibody waning in unvaccinated participants, a few substantial increases in antibody levels were observed. No reinfections were confirmed using qRT-PCR in self-collected saline gargle samples throughout the study, however one participant had a large (>8-fold) average increase in mean antibody levels (anti-S, RBD and N IgG) seven months following initial SARS-CoV-2 diagnosis, which may be explained by a second exposure to SARS-CoV-2. A second participant had 6-fold increase of anti-S and anti-RBD levels, but not anti-N IgG levels, suggesting they may also have been reexposed. Other detected increases in antibody levels were of much smaller magnitude and might be secondary to rising titers early in convalescence or be explained by technical variations rather than a biological mechanism.

### 4.2. Comparison with Literature

Previous studies have measured changes in SARS-CoV-2 antibody titers over time. Repeated exposure to SARS-CoV-2 antigens increases IgG titer, while antibodies generated from a single exposure wane overtime (42, 43). Following infection, SARS-CoV-2 specific antibody waning has been observed to decrease from the 8^th^ to 9^th^ week post symptoms onset, with detectable levels observed up to the end of the 12^th^ week (44). In those with multiple SARS-CoV-2 exposures or a hybrid immune response from infection and vaccination, decrease of antibody titers stops shortly after the secondary antigen exposure when stimulation of the memory B cell response produces additional antibodies (45). A strong correlation between total lymphocyte count and SARS-CoV-2 anti-S IgG provides evidence that an ongoing/active immune response provides better protection than a dormant one (44). Waning of SARS-CoV-2 antibodies differs by their paratope, anti-N IgG antibodies wane faster than anti-S. The difference in reactivity between anti-N to anti-S IgG was observed at the population level, anti-N seroprevalence underestimated the number of confirmed infections by 9-31% (46). Vaccination post SARSCoV-2 infection prevents waning of anti-S but has no effect on anti-N IgG (47). Hybrid immunity also benefits the breadth of the antibody mediated response, increasing the probability that existing antibodies are effective against the novel variants. Persons who were infected prior to receiving one of two doses of a COVID-19 vaccine had more somatic mutations and antibody production from the IGHV2-5; IGHJ4-1 germline which was not active in the vaccinated but uninfected (48, 49). Additionally, hybrid immunity produces greater total and neutralizing anti-S titers than natural infection or vaccination alone (45). Our study both supports and builds upon prior findings, as we show that SARS-CoV-2 IgG antibodies wane in SARS-CoV-2 infected people over time, with the rate of decline being greater for anti-N IgG than anti-S; vaccination post-infection boosts anti-S IgG titers and participants with hybrid immunity possess anti-S antibody levels which are not common in those infected but unvaccinated. Our calculated rate of antibody decline may be used to help estimate infection timing in seroprevalence studies.

### 4.3. Clinical and Epidemiological Interpretation

Our findings have important implications for clinical practice and public health guidelines as the pandemic progresses into its third year, novel viral variants continue to emerge, and vaccine doses are more widely distributed globally. Humoral immunity from natural infection wanes and vaccination with at least one dose of COVID-19 vaccine increases SARS-CoV-2 anti-S IgG titers immediately and over time. Therefore, we recommend that naturally infected individuals receive COVID-19 vaccination to increase protection from re-infection and severe disease and the duration of their humoral immune response against SARS-CoV-2. We demonstrate that a single dose of SARS-CoV-2 vaccine is effective in boosting anti-S antibody titers to high levels, which has implications in distribution of vaccine supplies in those countries with scarce access and low vaccination levels in the setting of high numbers of natural infection.

### 4.4 Strengths and Limitations

The strength of the described study stems from the prospective design, use of multiple serological tests, including the quantitative MSD option, and thorough analysis. A prospective cohort design offers several benefits, which allowed us to observe SARS-CoV-2 antibody dynamics over time with minimal bias. Recruiting participants post-infection but prior to vaccination delineated the sequence of temporal events, limiting the probability that any changes in antibody titers observed post-vaccination were due to causes other than the vaccine. Selection bias was minimized as the participants exposure and outcome status were not known when they were recruited into the study. At the beginning of the study, the measured covariates were exchangeable between participants who were unvaccinated or vaccinated during follow-up. Balance of the covariates allowed for estimation of the relationship between vaccination and anti-SARS-CoV-2 IgG antibody titer with minimal bias from confounding. Utilizing multiple serological tests strengthened our observations, as well as allows generalizability to study populations in many different laboratories. Statistical power was optimized by analysis with a mixed effects linear regression model, which accommodated multiple repeated measures per participant.

Limitations of the work include differential loss to follow up in the vaccinated and unvaccinated groups, a small sample size, and incomplete/missing survey responses. Unvaccinated participants were observed to have approximately one fewer visit than those who received a COVID-19 vaccine. Vaccines were not an originally planned intervention in the study and were made available in British Columbia on a stage roll-out basis about half-way through the study period. The difference in visit numbers between the vaccinated and unvaccinated groups is likely related to surveillance bias-those who receive a medical intervention are more open to clinical follow up than those who do not. Obtaining a larger sample size initially planned for the study was difficult due to low enrollment uptake, likely related to the social and economic stress of the pandemic on the public and geographic limitations on recruitment related to the availability of sample collection sites.

While the reached sample size was adequate for the primary analysis-which included repeated measures-enrolling additional participants would have allowed for a more precise estimate of the relationship between COVID-19 vaccination and endemic HCoVs anti-IgG titers. Although not significant, a trend was observed for increased average antibody titers for HCoV-HKU1 and HCoV-OC43 following vaccination which, like SARS-CoV-2, are both β-coronaviruses in contrast to decreases in the averages observed for both HCoV-NL63 and HCoV-229E which are α-coronaviruses. The study design may have also underestimated any association between existing endemic coronavirus IgG titers and COVID-19 vaccination as the sample was restricted to persons previously infected with SARS-CoV-2. COVID-19 has been shown to affect endemic coronavirus antibody levels and as such, the effect of vaccination should be observed in a cohort of SARS-CoV-2 naive persons prior to vaccination (50–52). The overall effect of SARS-CoV-2 vaccination and/or infection on the circulating antibodies against endemic HCoVs in the population may have implications for their seasonal epidemiology.

## 5. Conclusions

In summary, we report that single dose vaccination in a British Columbia-based cohort after natural infection significantly increases SARS-CoV-2 anti-S IgG titer by 1.63 log_10_ units and that vaccination increases the durability of high anti-S titers over time. Vaccination post-natural infection had no significant association with SARS-CoV-2 anti-N IgG titer; a non-significant trend towards higher anti-S IgG against the endemic β-HCoVs was observed. Our results provide support that vaccination is beneficial for achieving higher and more persistent SARS-CoV-2 anti-S IgG levels. We also report an estimated rate of decay of anti-N antibodies, which may be useful for ongoing population seroprevalence estimates. Future studies should examine the impact of infection following vaccination on antibody dynamics, as vaccine breakthrough infections with omicron or other variants continue to occur.

## Data Availability

The datasets generated during and/or analyzed during the current study are available from the corresponding author on reasonable request.

## Supplementary Materials

The following supporting information can be downloaded at: www.mdpi.com/xxx/s1, Table S1: Summary of serological test results; Figure S1: Exclusion criteria were applied to select an analytic data of n = 270 observations from k = 42 dependent participants (clusters).

## Author Contributions

“Conceptualization, **A.D.O, A.N.J, M.M** and **I.S**.; methodology, **A.M.N, A.C.M**,**E.F, M.S, A.H** and **S.M**; software, **A.D.O, A.M.N**; formal analysis, **A.D.O, A.M.N**; investigation, **A.D.O, I.S**; resources, **A.D.O, A.N.J, M.M, I.S, A.C.M, E.F, M.S, A.H** and **S.M**; data curation, **S.S, A.C.M** and **T.H**; writing—original draft preparation, **A.D.O, A.M.N**; writing—review and editing, **All Authors**; visualization, **A.D.O, A.M.N**; supervision, **A.N.J and I.S**; project administration, **A.C.M, E.F, A.H**, ; funding acquisition, **A.N.J, M.M** and **I.S**.. All authors have read and agreed to the published version of the manuscript.

## Institutional Review Board Statement

The study was conducted in accordance with the Declaration of Helsinki, and approved by the Institutional Review Board (or Ethics Committee) of The University of British Columbia (H20-01089 on 4/21/2020).

## Informed Consent Statement

Informed consent was obtained from all subjects involved in the study.

## Acknowledgments

We thank the dedicated staff at the BCCDC PHL for processing, testing, and sequencing clinical specimens. We also thank all the study participants who volunteered their time and specimens for this research. We also are thankful to all the staff who helped to collect and process samples for this study at Abbotsford Regional Hospital, Surrey Memorial Hospital and St. Paul’s Hospital. Notably, we thank the staff at BC Children’s Hospital for providing extra guidance and support to the researchers in conducting the study.

## References

1. Fung TS, Liu DiX. 2021. Similarities and Dissimilarities of COVID-19 and Other Coronavirus Diseases. https://doi.org/101146/annurev-micro-110520-023212 75:19–47.

2. Fehr AR, Perlman S. 2015. Coronaviruses: An Overview of Their Replication and Pathogenesis. Coronaviruses: Methods and Protocols 1282:1–23.

3. Huang AT, Garcia-Carreras B, Hitchings MDT, Yang B, Katzelnick LC, Rattigan SM, Borgert BA, Moreno CA, Solomon BD, Trimmer-Smith L, Etienne V, Rodriguez-Barraquer I, Lessler J, Salje H, Burke DS, Wesolowski A, Cummings DAT. 2020. A systematic review of antibody mediated immunity to coronaviruses: kinetics, correlates of protection, and association with severity. Nature Communications 2020 11:1 11:1–16.

4. Skowronski DM, Sekirov I, Sabaiduc S, Zou M, Morshed M, Lawrence D, Smolina K, Ahmed MA, Galanis E, Fraser MN, Singal M, Naus M, Patrick DM, Kaweski SE, Mill C, Reyes RC, Kelly MT, Levett PN, Petric M, Henry B, Krajden M. 2020. Low SARS-CoV-2 sero-prevalence based on anonymized residual sero-survey before and after first wave measures in British Columbia, Canada, March-May 2020. medRxiv 2020.07.13.20153148.

5. Long QX, Liu BZ, Deng HJ, Wu GC, Deng K, Chen YK, Liao P, Qiu JF, Lin Y, Cai XF, Wang DQ, Hu Y, Ren JH, Tang N, Xu YY, Yu LH, Mo Z, Gong F, Zhang XL, Tian WG, Hu L, Zhang XX, Xiang JL, D. HX, Liu HW, Lang CH, Luo XH, Wu SB, Cui XP, Zhou Z, Zhu MM, Wang J, Xue CJ, Li XF, Wang L, Li ZJ, Wang K, Niu CC, Yang QJ, Tang XJ, Zhang Y, Liu XM, Li JJ, Zhang DC, Zhang F, Liu P, Yuan J, Li Q, Hu JL, Chen J, Huang AL. 2020. Antibody responses to SARS-CoV-2 in patients with COVID-19. Nature Medicine 2020 26:6 26:845–848.

6. To KKW, Tsang OTY, Leung WS, Tam AR, Wu TC, Lung DC, Yip CCY, Cai JP, Chan JMC, Chik TSH, Lau DPL, Choi CYC, Chen LL, Chan WM, Chan KH, Ip JD, Ng ACK, Poon RWS, Luo CT, Cheng VCC, Chan JFW, Hung IFN, Chen Z, Chen H, Yuen KY. 2020. Temporal profiles of viral load in posterior oropharyngeal saliva samples and serum antibody responses during infection by SARS-CoV-2: an observational cohort study. Lancet Infect Dis 20:565–574.

7. Dorigatti I, Lavezzo E, Manuto L, Ciavarella C, Pacenti M, Boldrin C, Cattai M, Saluzzo F, Franchin E, Del Vecchio C, Caldart F, Castelli G, Nicoletti M, Nieddu E, Salvadoretti E, Labella B, Fava L, Guglielmo S, Fascina M, Grazioli M, Alvisi G, Vanuzzo MC, Zupo T, Calandrin R, Lisi V, Rossi L, Castagliuolo I, Merigliano S, Unwin HJT, Plebani M, Padoan A, Brazzale AR, Toppo S, Ferguson NM, Donnelly CA, Crisanti A. 2021. SARS-CoV-2 antibody dynamics and transmission from community-wide serological testing in the Italian municipality of Vo’. Nature Communications 2021 12:1 12:1–11.

8. Garcia-Beltran WF, Lam EC, Astudillo MG, Yang D, Miller TE, Feldman J, Hauser BM, Caradonna TM, Clayton KL, Nitido AD, Murali MR, Alter G, Charles RC, Dighe A, Branda JA, Lennerz JK, Lingwood D, Schmidt AG, Iafrate AJ, Balazs AB. 2021. COVID-19-neutralizing antibodies predict disease severity and survival. Cell 184:476-488.e11.

9. Noh JY, Kwak JE, Yang JS, Hwang SY, Yoon JG, Seong H, Hyun H, Lim CS, Yoon SY, Ryou J, Lee JY, Kim SS, Park SH, Cheong HJ, Kim WJ, Shin EC, Song JY. 2021. Longitudinal Assessment of Antisevere Acute Respiratory Syndrome Coronavirus 2 Immune Responses for Six Months Based on the Clinical Severity of Coronavirus Disease 2019. J Infect Dis 224:754–763.

10. Khoury DS, Cromer D, Reynaldi A, Schlub TE, Wheatley AK, Juno JA, Subbarao K, Kent SJ, Triccas JA, Davenport MP. 2021. Neutralizing antibody levels are highly predictive of immune protection from symptomatic SARS-CoV-2 infection. Nature Medicine 2021 27:7 27:1205–1211.

11. Dolscheid-Pommerich R, Bartok E, Renn M, Kümmerer BM, Schulte B, Schmithausen RM, Stoffel-Wagner B, Streeck H, Saschenbrecker S, Steinhagen K, Hartmann G. 2022. Correlation between a quantitative anti-SARS-CoV-2 IgG ELISA and neutralization activity. J Med Virol 94:388–392.

12. Garcia-Beltran WF, Lam EC, Astudillo MG, Yang D, Miller TE, Feldman J, Hauser BM, Caradonna TM, Clayton KL, Nitido AD, Murali MR, Alter G, Charles RC, Dighe A, Branda JA, Lennerz JK, Lingwood D, Schmidt AG, Iafrate AJ, Balazs AB. 2021. COVID-19-neutralizing antibodies predict disease severity and survival. Cell 184:476-488.e11.

13. Dogan M, Kozhaya L, Placek L, Gunter C, Yigit M, Hardy R, Plassmeyer M, Coatney P, Lillard K, Bukhari Z, Kleinberg M, Hayes C, Arditi M, Klapper E, Merin N, Liang BTT, Gupta R, Alpan O, Unutmaz D. 2021. SARS-CoV-2 specific antibody and neutralization assays reveal the wide range of the humoral immune response to virus. Communications Biology 2021 4:1 4:1–13.

14. Solís Arce JS, Warren SS, Meriggi NF, Scacco A, McMurry N, Voors M, Syunyaev G, Malik AA, Aboutajdine S, Adeojo O, Anigo D, Armand A, Asad S, Atyera M, Augsburg B, Awasthi M, Ayesiga GE, Bancalari A, Björkman Nyqvist M, Borisova E, Bosancianu CM, Cabra García MR, Cheema A, Collins E, Cuccaro F, Farooqi AZ, Fatima T, Fracchia M, Galindo Soria ML, Guariso A, Hasanain A, Jaramillo S, Kallon S, Kamwesigye A, Kharel A, Kreps S, Levine M, Littman R, Malik M, Manirabaruta G, Mfura JLH, Momoh F, Mucauque A, Mussa I, Nsabimana JA, Obara I, Otálora MJ, Ouédraogo BW, Pare TB, Platas MR, Polanco L, Qureshi JA, Raheem M, Ramakrishna V, Rendrá I, Shah T, Shaked SE, Shapiro JN, Svensson J, Tariq A, Tchibozo AM, Tiwana HA, Trivedi B, Vernot C, Vicente PC, Weissinger LB, Zafar B, Zhang B, Karlan D, Callen M, Teachout M, Humphreys M, Mobarak AM, Omer SB. 2021. COVID-19 vaccine acceptance and hesitancy in low- and middle-income countries. Nature Medicine 2021 27:8 27:1385–1394.

15. Padma T V. 2021. COVID vaccines to reach poorest countries in 2023 - despite recent pledges. Nature 595:342–343.

16. Mathieu E, Ritchie H, Ortiz-Ospina E, Roser M, Hasell J, Appel C, Giattino C, Rodés-Guirao L. 2021. A global database of COVID-19 vaccinations. Nat Hum Behav 5:947–953.

17. Tao K, Tzou PL, Nouhin J, Gupta RK, de Oliveira T, Kosakovsky Pond SL, Fera D, Shafer RW. 2021. The biological and clinical significance of emerging SARS-CoV-2 variants. Nature Reviews Genetics 2021 22:12 22:757–773.

18. Planas D, Saunders N, Maes P, Guivel-Benhassine F, Planchais C, Buchrieser J, Bolland W-H, Porrot F, Staropoli I, Lemoine F, Péré H, Veyer D, Puech J, Rodary J, Baele G, Dellicour S, Raymenants J, Gorissen S, Geenen C, Vanmechelen B, Wawina-Bokalanga T, Martí-Carreras J, Cuypers L, Sève A, Hocqueloux L, Prazuck T, Rey F, Simon-Loriere E, Bruel T, Mouquet H, André E, Schwartz O. 2021. Considerable escape of SARS-CoV-2 Omicron to antibody neutralization. Nature 2021 1–7.

19. Bernal JL, Andrews N, Gower C, Gallagher E, Simmons R, Thelwall S, Stowe J, Tessier E, Groves N, Dabrera G, Myers R, Campbell CNJ, Amirthalingam G, Edmunds M, Zambon M, Brown KE, Hopkins S, Chand M, Ramsay M. 2021. Effectiveness of Covid-19 Vaccines against the B.1.617.2 (Delta) Variant. NEJM 385:585–594.

20. Sokal A, Barba-Spaeth G, Fernández I, Broketa M, Azzaoui I, de La Selle A, Vandenberghe A, Fourati S, Roeser A, Meola A, Bouvier-Alias M, Crickx E, Languille L, Michel M, Godeau B, Gallien S, Melica G, Nguyen Y, Zarrouk V, Canoui-Poitrine F, Pirenne F, Mégret J, Pawlotsky JM, Fillatreau S, Bruhns P, Rey FA, Weill JC, Reynaud CA, Chappert P, Mahévas M. 2021. mRNA vaccination of naive and COVID-19-recovered individuals elicits potent memory B cells that recognize SARS-CoV-2 variants. Immunity 54:2893-2907.e5.

21. Goel RR, Painter MM, Apostolidis SA, Mathew D, Meng W, Rosenfeld AM, Lundgreen KA, Reynaldi A, Khoury DS, Pattekar A, Gouma S, Kuri-Cervantes L, Hicks P, Dysinger S, Hicks A, Sharma H, Herring S, Korte S, Baxter AE, Oldridge DA, Giles JR, Weirick ME, McAllister CM, Awofolaju M, Tanenbaum N, Drapeau EM, Dougherty J, Long S, D’Andrea K, Hamilton JT, McLaughlin M, Williams JC, Adamski S, Kuthuru O, Frank I, Betts MR, Vella LA, Grifoni A, Weiskopf D, Sette A, Hensley SE, Davenport MP, Bates P, Luning Prak ET, Greenplate AR, Wherry EJ. 2021. mRNA vaccines induce durable immune memory to SARS-CoV-2 and variants of concern. Science (1979) 374.

22. Stamatatos L, Czartoski J, Wan Y-H, Homad LJ, Rubin V, Glantz H, Neradilek M, Seydoux E, Jennewein MF, Maccamy AJ, Feng J, Mize G, De Rosa SC, Finzi A, Lemos MP, Cohen KW, Moodie Z, Mcelrath MJ, Mcguire AT. 2021. mRNA vaccination boosts cross-variant neutralizing antibodies elicited by SARS-CoV-2 infection. Science (1979) 372:1413.

23. Harris PA, Taylor R, Thielke R, Payne J, Gonzalez N, Conde JG. 2009. Research electronic data capture (REDCap)—A metadata-driven methodology and workflow process for providing translational research informatics support. J Biomed Inform 42:377–381.

24. British Columbia Centre for Disease Control. 2021. Weekly Update on Variants of Concern. BC COVID-19 Data. Vancouver. http://www.bccdc.ca/Health-Info-Site/Documents/VoC/VoC_weekly_09172021.pdf. Retrieved 15 September 2022.

25. R Core Team. 2013. R: A language and environment for statistical computing. 3.6.3-"Holding the Windsock". R Foundation for Statistical Computing, Vienna, Austria.

26. Goldfarb DM, Tilley P, Al-Rawahi GN, Srigley JA, Ford G, Pedersen H, Pabbi A, Hannam-Clark S, Charles M, Dittrick M, Gadkar VJ, Pernica JM, Hoanga LMN. 2021. Self-Collected Saline Gargle Samples as an Alternative to Health Care Worker-Collected Nasopharyngeal Swabs for COVID-19 Diagnosis in Outpatients. J Clin Microbiol 59.

27. Hickman R, Nguyen J, Lee TD, Tyson JR, Azana R, Tsang F, Hoang L, Prystajecky N. 2022. Rapid, High-Throughput, Cost Effective Whole Genome Sequencing of SARS-CoV-2 Using a Condensed One Hour Library Preparation of the Illumina DNA Prep Kit. medRxiv 2022.02.07.22269672.

28. Sekirov I, Barakauskas VE, Simons J, Cook D, Bates B, Burns L, Masud S, Charles M, McLennan M, Mak A, Chahil N, Vijh R, Hayden A, Goldfarb D, Levett PN, Krajden M, Morshed M. 2021. SARS-CoV-2 serology: Validation of high-throughput chemiluminescent immunoassay (CLIA) platforms and a field study in British Columbia. J Clin Virol 142.

29. Stein DR, Osiowy C, Gretchen A, Thorlacius L, Fudge D, Lang A, Sekirov I, Morshed M, Levett PN, Tran V, Kus J v., Gubbay J, Mohan V, Charlton C, Kanji JN, Tipples G, Serhir B, Therrien C, Roger M, Jiao L, Zahariadis G, Needle R, Gilbert L, Desnoyers G, Garceau R, Bouhtiauy I, Longtin J, El-Gabalawy N, Dibernardo A, Lindsay LR, Drebot M. 2021. Evaluation of commercial SARS-CoV-2 serological assays in Canadian public health laboratories. Diagn Microbiol Infect Dis 101.

30. Li FF, Liu A, Gibbs E, Tanunliong G, Marquez AC, Gantt S, Frykman H, Krajden M, Morshed M, Prystajecky NA, Cashman N, Sekirov I, Jassem AN. 2022. A novel multiplex electrochemiluminescent immunoassay for detection and quantification of anti-SARS-CoV-2 IgG and anti-seasonal endemic human coronavirus IgG. J Clin Virol 146:105050.

31. Tanunliong G, Liu AC, Kaweski S, Irvine M, Reyes RC, Purych D, Krajden M, Morshed M, Sekirov I, Gantt S, Skowronski DM, Jassem AN. 2022. Age-Associated Seroprevalence of Coronavirus Antibodies: Population-Based Serosurveys in 2013 and 2020, British Columbia, Canada. Front Immunol 13.

32. Cohen J. 1992. A power primer. Psychol Bull 112:155–159.

33. Galecki A, Burzykowski T. 2013. Linear Mixed-Effects Model 245–273.

34. Vanderweele TJ, Shpitser I. 2011. A New Criterion for Confounder Selection. Biometrics 67:1406–1413.

35. Shoukri MM, Donner A, El-Dali A. 2013. Covariate-adjusted confidence interval for the intraclass correlation coefficient. Contemp Clin Trials 36:244–253.

36. Bozdogan H. 1987. Model selection and Akaike’s Information Criterion (AIC): The general theory and its analytical extensions. Psychometrika 1987 52:3 52:345–370.

37. Satagopan JM, Ben-Porat L, Berwick M, Robson M, Kutler D, Auerbach AD. 2004. A note on competing risks in survival data analysis. British Journal of Cancer 2004 91:7 91:1229–1235.

38. Bland JM, Altman DG. 2004. The logrank test. BMJ 328:1073.

39. BC Centre for Disease Control. 2022. BCCDC COVID-19 Situational Report - Week 2: January 09-January 15, 2022.

40. Hadfield J, Megill C, Bell SM, Huddleston J, Potter B, Callender C, Sagulenko P, Bedford T, Neher RA. 2018. NextStrain: Real-time tracking of pathogen evolution. Bioinformatics 34:4121–4123.

41. Torbati E, Krause KL, Ussher JE. 2021. The Immune Response to SARS-CoV-2 and Variants of Concern. Viruses 13:1911.

42. Lau CS, Phua SK, Liang YL, Lin M, Oh H, Aw TC. 2022. SARS-CoV-2 Spike and Neutralizing Antibody Kinetics 90 Days after Three Doses of BNT162b2 mRNA COVID-19 Vaccine in Singapore. Vaccines 2022, Vol 10, Page 331 10:331.

43. Levin EG, Lustig Y, Cohen C, Fluss R, Indenbaum V, Amit S, Doolman R, Asraf K, Mendelson E, Ziv A, Rubin C, Freedman L, Kreiss Y, Regev-Yochay G. 2021. Waning Immune Humoral Response to BNT162b2 Covid-19 Vaccine over 6 Months. New England Journal of Medicine 385:e84.

44. Li K, Huang B, Wu M, Zhong A, Li L, Cai Y, Wang Z, Wu L, Zhu M, Li J, Wang Z, Wu W, Li W, Bosco B, Gan Z, Qiao Q, Wu J, Wang Q, Wang S, Xia X. 2020. Dynamic changes in anti-SARS-CoV-2 antibodies during SARS-CoV-2 infection and recovery from COVID-19. Nat Commun 11:6044.

45. Bates TA, McBride SK, Leier HC, Guzman G, Lyski ZL, Schoen D, Winders B, Lee J-Y, Lee DX, Messer WB, Curlin ME, Tafesse FG. 2022. Vaccination before or after SARS-CoV-2 infection leads to robust humoral response and antibodies that effectively neutralize variants. Sci Immunol https://doi.org/10.1126/sciimmunol.abn8014.

46. Fenwick C, Croxatto A, Coste AT, Pojer F, André C, Pellaton C, Farina A, Campos J, Hacker D, Lau K, Bosch B-J, Gonseth Nussle S, Bochud M, D’Acremont V, Trono D, Greub G, Pantaleo G. 2021. Changes in SARS-CoV-2 Spike versus Nucleoprotein Antibody Responses Impact the Estimates of Infections in Population-Based Seroprevalence Studies. J Virol 95.

47. Jalkanen P, Kolehmainen P, Häkkinen HK, Huttunen M, Tähtinen PA, Lundberg R, Maljanen S, Reinholm A, Tauriainen S, Pakkanen SH, Levonen I, Nousiainen A, Miller T, Välimaa H, Ivaska L, Pasternack A, Naves R, Ritvos O, Österlund P, Kuivanen S, Smura T, Hepojoki J, Vapalahti O, Lempainen J, Kakkola L, Kantele A, Julkunen I. 2021. COVID-19 mRNA vaccine induced antibody responses against three SARS-CoV-2 variants. Nat Commun 12:3991.

48. Andreano E, Paciello I, Piccini G, Manganaro N, Pileri P, Hyseni I, Leonardi M, Pantano E, Abbiento V, Benincasa L, Giglioli G, de Santi C, Fabbiani M, Rancan I, Tumbarello M, Montagnani F, Sala C, Montomoli E, Rappuoli R. 2021. Hybrid immunity improves B cells and antibodies against SARS-CoV-2 variants. Nature 600:530–535.

49. Wang Z, Muecksch F, Schaefer-Babajew D, Finkin S, Viant C, Gaebler C, Hoffmann H-H, Barnes CO, Cipolla M, Ramos V, Oliveira TY, Cho A, Schmidt F, da Silva J, Bednarski E, Aguado L, Yee J, Daga M, Turroja M, Millard KG, Jankovic M, Gazumyan A, Zhao Z, Rice CM, Bieniasz PD, Caskey M, Hatziioannou T, Nussenzweig MC. 2021. Naturally enhanced neutralizing breadth against SARS-CoV-2 one year after infection. Nature 595:426–431.

50. Guo L, Wang Y, Kang L, Hu Y, Wang L, Zhong J, Chen H, Ren L, Gu X, Wang G, Wang C, Dong X, Wu C, Han L, Wang Y, Fan G, Zou X, Li H, Xu J, Jin Q, Cao B, Wang J. 2021. Cross-reactive antibody against human coronavirus OC43 spike protein correlates with disease severity in COVID-19 patients: a retrospective study. Emerg Microbes Infect 10:664.

51. Lin CY, Wolf J, Brice DC, Sun Y, Locke M, Cherry S, Castellaw AH, Wehenkel M, Crawford JC, Zarnitsyna VI, Duque D, Allison KJ, Allen EK, Brown SA, Mandarano AH, Estepp JH, Gaur AH, Hoffman JM, Mori T, Tuomanen EI, Webby RJ, Hakim H, Hayden RT, Hijano DR, Awad W, Bajracharya R, Clark BL, Cortez V, Dallas RH, Fabrizio T, Freiden P, Gowen A, Hodges J, Kirk AM, Roubidoux EK, Mettelman RC, Russell-Bell J, Souquette A, Sparks J, van de Velde LA, Vazquez-Pagan A, Whitt K, Wilson TL, Wittman DE, Wohlgemuth N, Wu G, Taylor C, Molina-Paris C, Schultz-Cherry S, Tang L, Thomas PG, McGargill MA. 2022. Pre-existing humoral immunity to human common cold coronaviruses negatively impacts the protective SARS-CoV-2 antibody response. Cell Host Microbe 30:83-96.e4.

52. Anderson EM, Goodwin EC, Verma A, Arevalo CP, Bolton MJ, Weirick ME, Gouma S, McAllister CM, Christensen SR, Weaver JE, Hicks P, Manzoni TB, Oniyide O, Ramage H, Mathew D, Baxter AE, Oldridge DA, Greenplate AR, Wu JE, Alanio C, D’Andrea K, Kuthuru O, Dougherty J, Pattekar A, Kim J, Han N, Apostolidis SA, Huang AC, Vella LA, Kuri-Cervantes L, Pampena MB, Betts MR, Wherry EJ, Meyer NJ, Cherry S, Bates P, Rader DJ, Hensley SE. 2021. Seasonal human coronavirus antibodies are boosted upon SARS-CoV-2 infection but not associated with protection. Cell 184:1858.

