## Supplementary material for "Characterizing Longitudinal Antibody Responses in Recovered Individuals Following COVID-19 Infection and Single-Dose Vaccination in British Columbia, Canada: a Prospective Cohort Study": Table S1 and Figure S1

**Supplemental Materials:

Table S1.** Summary of serological test results.

| **Test** | **No. samples**  **tested** | **No. samples**  **positive (%)** | **No. (%) positive**  **<6 months** | **No.% positive**  **>6 months** | ***P-Value** |
| --- | --- | --- | --- | --- | --- |
| Total anti-RBD^$^(Siemens) | 340 | 324 (95%) | (191/196) 97% | (133/144) 92% | 0.05 |
| Anti-N IgG (Abbott) | 335 | 158 (47%) | (140/194) 72% | (18/141) 13% | <0.001 |
| Total anti-S (Ortho) | 181 | 181 (100%) | (56/56) 100% | (125/125) 100% | -- |
| Anti-RBD^$^ IgG (MSD) | 339 | 324 (96%) | (189/196) 96% | (135/143) 94% | 0.53 |
| Anti-N IgG (MSD) | 339 | 196 (58%) | (149/196) 76% | (47/143) 33% | <0.001 |
| Anti-S IgG (MSD) | 339 | 303 (89%) | (181/196) 92% | (122/143) 85% | 0.06 |

^$^ 2 samples negative by anti-RBD (Siemens) were not available for Abbott or Ortho testing
* P-Values are reported for a Chi-square test (χ²test) for two independent proportions.

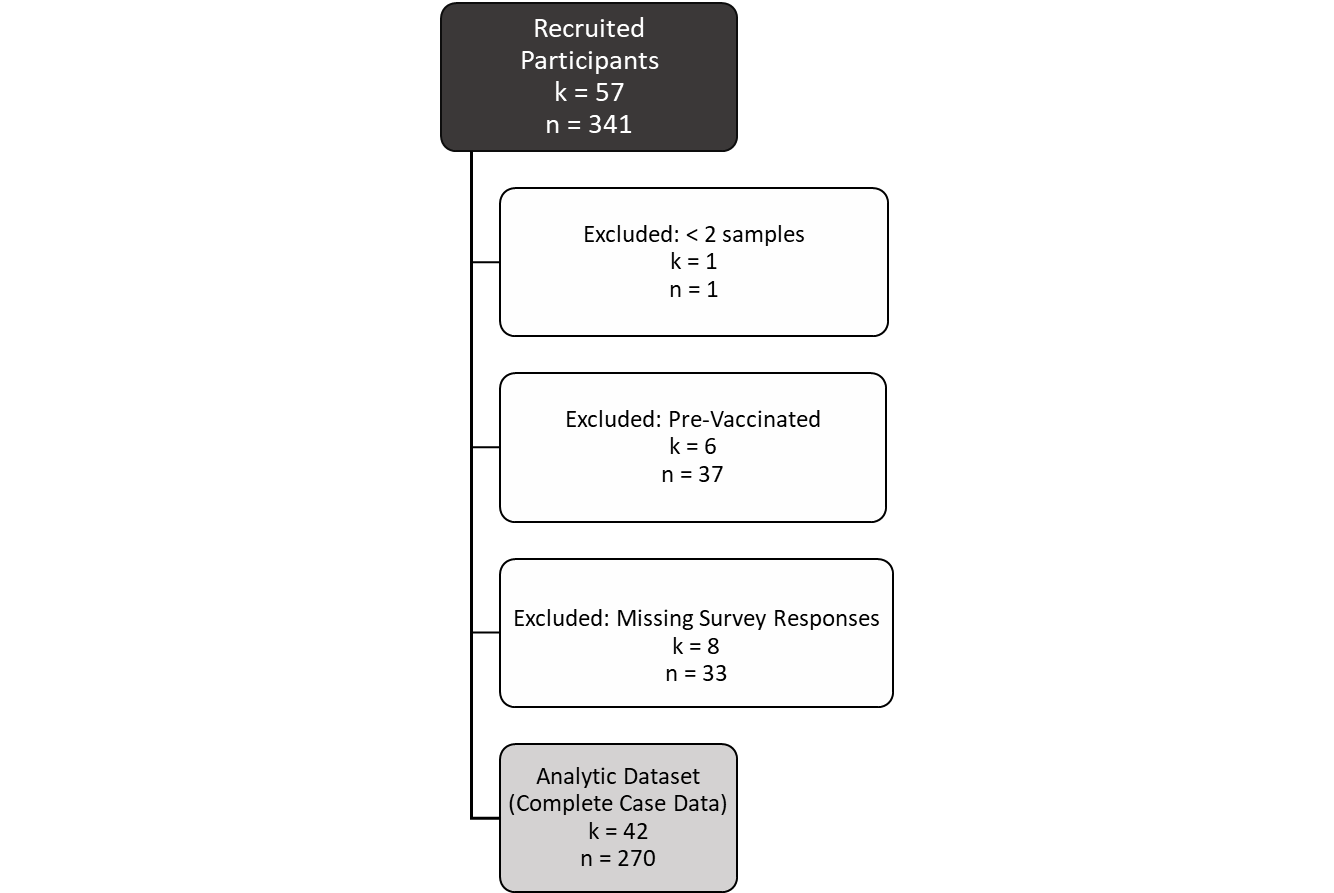

**Figure S1.** Exclusion criteria were applied to select an analytic data of n = 270 observations from k = 42 dependent participants (clusters).
